# Automated Western immunoblotting detection of anti-SARS-CoV-2 serum antibodies

**DOI:** 10.1101/2020.11.26.20238733

**Authors:** S. Edouard, R. Jaafar, N. Orain, P. Parola, P. Colson, B. La Scola, P-E. Fournier, D. Raoult, M. Drancourt

## Abstract

ELISA and chemiluminescence serological assays for COVID-19 are currently incorporating only one or two SARS-CoV-2 antigens. We developed an automated Western immunoblotting as a complementary serologic assay for COVID-19. The Jess™ Simple Western system, an automated capillary-based assay was used, incorporating an inactivated SARS-CoV-2 lineage 20a strain as antigen, and IgT detection. In total, 602 sera were tested including 223 from RT-PCR-confirmed COVID-19 patients, 76 from patients diagnosed with seasonal HCoVs and 303 from coronavirus-negative control sera. We also compared this assay with the EUROIMMUN® SARS-CoV-2 IgG ELISA kit. Among 223 sera obtained from RT-PCR-confirmed COVID-19 patients, 180/223 (81%) exhibited reactivity against the nucleocapsid and 70/223 (31%) against the spike protein. Nucleocapsid reactivity was further detected in 9/76 (14%) samples collected from patients diagnosed with seasonal HCoVs and in 15/303 (5%) coronavirus-negative control samples. In the subset of sera collected more than 2 weeks after the onset of symptoms, the sensitivity was 94% and the specificity 93%, the latter value probably reflecting cross-reactivity of SARS-CoV-2 with other coronaviruses. The automated Western immunoblotting presented a substantial agreement (90%) with the compared ELISA (Cohen’s Kappa=0.64). Automated Western immunoblotting may be used as a second line test to monitor exposition of people to HCoVs including SARS-CoV-2.

## 1. INTRODUCTION

To date, seven coronaviruses have been reported as human pathogens, including four seasonal coronaviruses (*Alphacoronavirus* 229E and NL63 and *Betacoronavirus* HKU1 and OC43) here referred to as HCoVs, which are associated to mild-to-severe upper and lower respiratory tract infections (1). Two other betacoronaviruses that caused severe acute respiratory syndrome in 2002 in China (SARS-CoV) and the Middle East Respiratory Syndrome in 2012 in Saudi Arabia (MERS-CoV) (2); and the *Betacoronavirus* SARS-CoV-2 that is the agent of the COVID-19 pandemic has been demonstrated to infect a variety of animals and humans (3). The latter is phylogenetically closely related to HCoV-HKU1 and presents a high sequence homology with SARS-CoV (2).

Serological assays used to explore exposition to seasonal HCoVs have previously indicated cross-immunity between all coronaviruses (4–6). SARS-CoV-2 exhibits several antigens eliciting a serological response in COVID-19 patients, including spike glycoprotein, its N-terminal (S1) and C-terminal (S2) subunits as well as nucleocapsid (7). Most of routinely used serological COVID-19 assays incorporated only one recombinant protein (8–10). Second generation assays are combining two antigens to increase sensitivity and mostly specificity (7, 11).

We developed an automated Western immunoblotting (AWB) assay in order to characterize serological responses to SARS-CoV-2 and the potential cross-reactivity with HCoVs.

## 2. PATIENTS AND METHODS

### Serum sample collections

A first set of 27 serum samples from 27 different patients with RT-PCR-documented COVID-19 (12), collected at least 2 weeks after the onset of symptoms were incorporated as a positive control group. All of them presented IgG titer ≥ 1:100 using in-house indirect immunofluorescence assay (IFA) (13). Of these, 16 serum samples were used for conventional immunoblotting including 3 samples exhibiting low (1:200), moderate (1:800) and high (1:3,200) IgG titers using IFA that were used to fix optimal conditions to be used for AWB (antigen, serum and secondary antibodies concentrations). One serum collected in 2018, before the onset of COVID-19 (negative RT-PCR for HCoVs on homologous respiratory specimen) was included as negative control.

As for AWB, 223 serum samples (including the 27 serum samples described above) collected from 223 different RT-PCR-confirmed COVID-19 patients were incorporated as a positive control group. Twenty-seven of these sera were tested for antibodies to the recombinant S1 protein by EUROIMMUN® SARS-CoV-2 IgG ELISA (Euroimmun, Bussy Saint-Martin, France) performed using the Elispeed DUO system (Euroimmun) according to the manufacturer’s recommendations. The ratio (AUC sample/AUC calibrator) was interpreted as follows: <0.8 negative; ≥0.8 to <1.0 undetermined; ≥1.1 positive. We considered undetermined results as negative for statistical analyses. A negative control group (37 serum samples) consisted of (i) 10 serum sampled obtained less than 5 days after the onset of symptoms) in patients presenting high viral loads of SARS-CoV-2 (Ct values < 20); (ii) 14 sera from asymptomatic healthcare workers largely exposed to the virus but exhibiting negative results for RT-PCR and serology by IFA for SARS-CoV-2 during follow-up; and (iii) 13 sera from patients collected in 2019 before the pandemic and harbouring negative RT-PCR results for the 4 HCoVs in their nasopharyngeal specimens. These 37 serum samples were also all tested by ELISA. A third group of 76 serum samples was retrieved from patients diagnosed with seasonal coronavirus infections (HCoV-NL63 (n=19), HCoV-OC43 (n=21), HCoV-229E (n=8) and HCoV-HKU1 (n=28)) and were collected at least 2 weeks after the diagnosis, of which 45 were also tested by ELISA. A fourth group of 266 sera was collected from children and adults admitted in surgery departments (n=145) and other medical units (=121) before the pandemic, of which 88 serum samples were also tested by ELISA; their HCoVs status was unknown. Altogether, 197 sera tested by ELISA, included 27 sera from COVID-19-positive patients and 170 from COVID-19-negative patients. All sera were retrospectively tested and no sample was collected specifically for this study which was approved by our institution’s ethics committee under No.2020-024.

### Virus growth, purification and concentration

The SARS-CoV-2 IHUMI2 strain (lineage 20a) was used as antigen as previously described (13). One liter of infected cells was collected and clarified by centrifugation at 700 x g for 10 min and by filtering the supernatant through a 0.45-µm pore-sized filter and further a 0.2-µm pore-sized filter. Virions were then aggregated by overnight precipitation at 4°C with 10% polyethylene glycol 8000 white flake type (PEG-8000, BioUltra, SIGMA-ALDRICH, USA) and 2.2% crystalline NaCl, with gentle swirling. Precipitated virus particles were then centrifuged at 10,000 x g for 30 min using a SORVALL Evolution centrifuge with SLA-3000 Recent 1 fixed angle rotor pre-cooled at 4°C (Kendro Laboratory Products, Newtown, USA). The pellet was resuspended with HEPES-saline (0.9% NaCl, 10 mL of 1 M HEPES, 990 mL purified water) previously vacuum-sterilized through a 0.2-µm pore size membrane; swirled in the cold HEPES-saline until dissolution to avoid using pipetting as it may hurts viral spikes at this step. The resuspended pellet was then treated with a 30% sucrose cushion in 25 x 89 mm centrifuge tubes (Ultra-Clear, BECKMAN COULTER, CA, USA). Final purification was achieved by ultracentrifugation at 100,000 x g for 90 min at 4°C followed by two 30-min washes with HBSS using SORVALL Discovery 90SE with Surespin 630 rotor (Kendro Laboratory Products). The final pellet was resuspended in 400 µL of HEPES-buffered saline and heat-inactivated at 65°C for 1h.

### Conventional Western immunoblotting

SARS-CoV-2 antigens diluted to 0.5 mg/mL were mixed (v/v) with 2X Laemmli Sample Buffer (Bio-Rad, Hercules, CA, USA) before a heating step of 5 min at 95°C. This preparation and a ladder were dispensed in wells shaped in a 5 % polyacrylamide stacking gel. The protein separation was then performed in a 10 % polyacrylamide separating gel with a Mini Trans-blot cell device (Bio-Rad) at 160 V for 90 min. After transferring proteins from the gel to a 0.45 μm-pore size nitrocellulose membrane (Bio-Rad) at 100 V and 15°C for 90 min, the membrane was left at 4°C overnight with 5% non-fat milk powder in TBS buffer. Blocked strips were incubated with sera diluted at 1:50 for 60 min. Three washes of 10 min were performed before a 90-min incubation of the strips with goat peroxidase-conjugated anti-human IgG/ IgM/IgA (Jackson ImmunoResearch, Ely, UK) diluted 1:1000. Three washes of 10 min with TBS buffer were made. Strips were put in contact with ECL Western Blotting Substrate (Promega, Madison, USA) and the reaction with secondary antibody peroxydases was revelated with a Fusion Fx chemiluminescence imaging system and analysed with the Fusion software (Vilber, Marne-la-Vallée, France).

### Automated Western immunoblotting

The Jess™ Simple Western system (ProteinSimple, San Jose CA, USA,) is an automated capillary-based size separation and nano-immunoassay system. To quantify the absolute serological response to viral antigens, we followed the manufacturer’s standard method for 12-230-kDa Jess separation module (SM-W004). The SARS-CoV-2 antigen (1 µg/µL) was mixed with 0.1X Sample buffer and Fluorescent 5X Master mix (ProteinSimple) to achieve a final concentration of 0.25 µg/µL in the presence of fluorescent molecular weight markers and 400 mM dithiothreitol (ProteinSimple). This preparation was denatured at 95°C for 5 minutes. Ladder (12-230-kDa PS-ST01EZ) and SARS-CoV-2 proteins were separated in capillaries as they migrated through a separation matrix at 375 volts. A ProteinSimple proprietary photoactivated capture chemistry was used to immobilize separated viral proteins on the capillaries. Patients sera diluted at a 1:2 were added and incubated for 60 min. After a wash step, goat HRP-conjugated anti-human IgG/IgM/IgA antibodies (Jackson ImmunoResearch) diluted 1:500 was added for 30 min. The chemiluminescent revelation was established with peroxyde/luminol-S (ProteinSimple). Digital image of chemiluminescence of the capillary was captured with Compass Simple Western software (version 4.1.0, Protein Simple) that calculated automatically heights (chemiluminescence intensity), area and signal/noise ratio. Results could be visualized as electropherograms representing peak of chemiluminescence intensity and as lane view from signal of chemiluminescence detected in the capillary. An internal system control was included in each run.

### Statistical analysis

ROC curves were performed using XL stat. The agreement rate and Cohen’s Kappa value were determined for agreement between ELISA and AWB. For data comparisons and statistical analyses, the Fisher’s exact test, Chi-squared test, Mann-Witney test and standard statistical software (GraphPad Prism v7) were used. A p-value < 0.05 was considered statistically significant.

## 3. RESULTS

### Fixing automated Western immunoblotting parameters

Protein profiles of the purified SARS-CoV-2 antigen and uninfected Vero cells were verified on silver-stained 2-D gel. As expected, the viral specific and major dominant proteins were N, S, S1 and S2 proteins at 42, 170, 110 and 90 kDa, respectively. All 16 serum samples collected from 16 different COVID-19 patients exhibited reactivity against the nucleocapsid and spike proteins. Parameter optimization to translate these data on AWB included an antigen concentration of 0.25 µg/µL, a serum dilution at 1:2 and a secondary antibody dilution of 1:500 (data not shown). In these conditions, AWB of positive serum samples yielded a prominent 56-kDa band interpreted as the nucleocapsid and a 217-kDa band interpreted as the spike protein (Figure S1). Higher molecular weight values observed with AWB than with SDS-PAGE were due to the different composition of gel in the capillaries. In total, the 16 sera from COVID-19 patients tested with conventional and AWB gave similar results except that AWB failed to detect the spike protein in one sample. Further, AWB yielded significant higher S/N ratio, pick height and area under curve for the nucleocapsid (*p* < 0.0001) and spike proteins (*p* < 0.0001) in the 27 serum samples from COVID-19 patients than in 37 serum samples collected in negative control patients (Table 1). The S/N ratio presented higher Youden Index for nucleocapsid detection, being therefore interpreted as the most pertinent parameter to interpret AWB results. Optimal threshold for the S/N ratio of 110.4 conferred a 96.3% sensitivity and 94.6% specificity for the nucleocapsid detection. Determination of a cut-off to interpret results of spike protein was not useful and could be based only on presence/absence of signal with sensitivity to 66.7% and 100% specificity (Table 2). Therefore, we further used the presence of antibodies to the nucleocapsid with S/N ratio ≥ 110.4 and/or to the spike protein, as criteria to define a positive AWB.

**Table 1.**
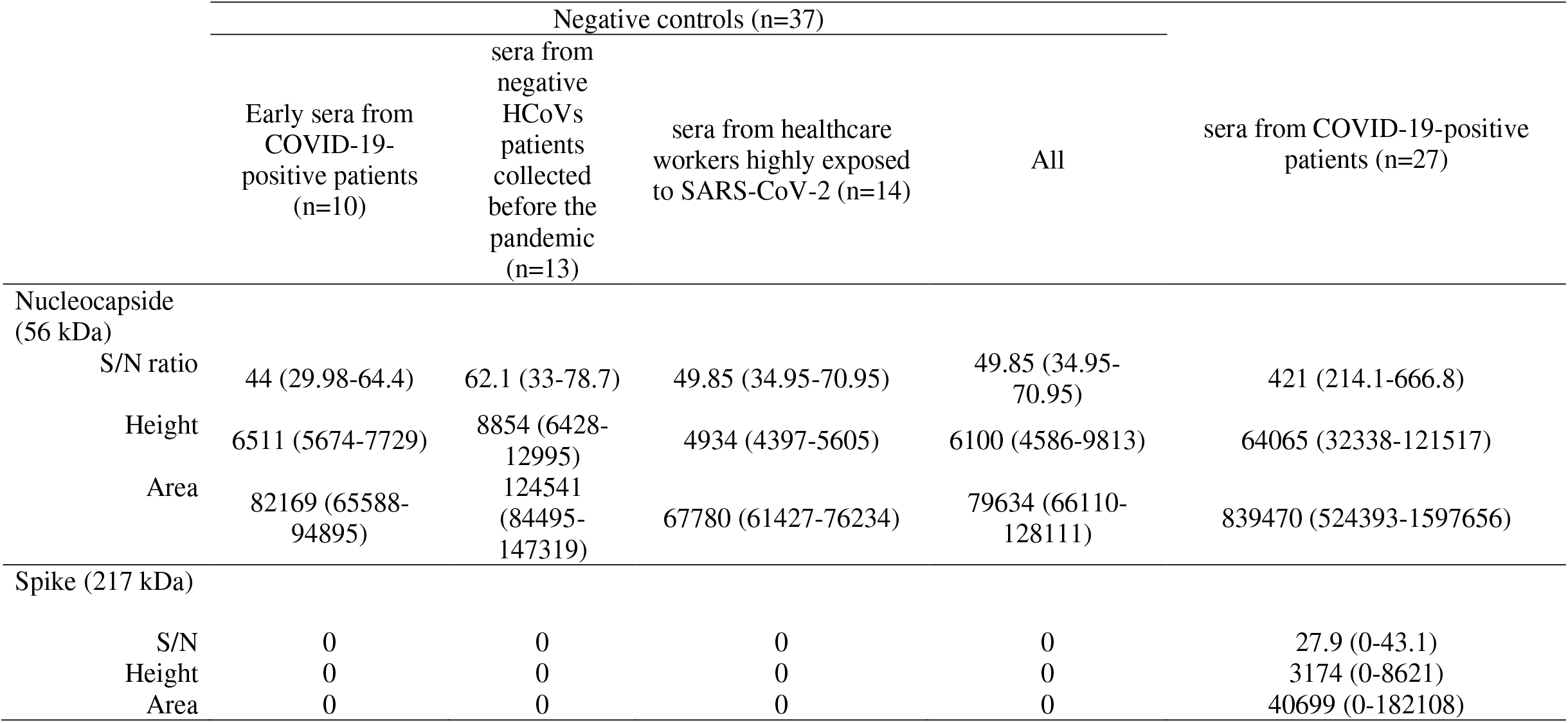
**Automated Western immunoblotting results of 27 sera from COVID-19-positive patients and 37 sera from negative controls used to fix automated Western immunoblotting parameters. (**Results expressed as median with 25 and 75 percentile).

**Table 2.** **Sensitivity and specificity of automated Western immunoblotting determined with 27 sera from COVID-19-positive patients and 37 sera from non COVID-19 patients**.

| n=64 | AUC | Youden index | Optimal Cut-off value | Sensitivity (%) | Specificity (%) | True positive (nb) | True negative (nb) | False positive (nb) | False negative (nb) |
| --- | --- | --- | --- | --- | --- | --- | --- | --- | --- |
| Nucleocapside |  |  |  |  |  |  |  |  |  |
| S/N | 0.975 | 0.907 | 110.4 | 96.3 | 94.6 | 26 | 35 | 2 | 1 |
| Height | 0.982 | 0.885 | 24922 | 88.9 | 100 | 24 | 37 | 0 | 3 |
| Area | 0.970 | 0.885 | 287005 | 88.9 | 100 | 24 | 37 | 0 | 3 |
| Spike |  |  |  |  |  |  |  |  |  |
| S/N | 0.833 | 0.667 | 0 | 66.7 | 100 | 18 | 37 | 0 | 9 |
| Height | 0.833 | 0.667 | 0 | 66.7 | 100 | 18 | 37 | 0 | 9 |
| Area | 0.833 | 0.667 | 0 | 66.7 | 100 | 18 | 37 | 0 | 9 |

AUC= area under the curve.

### Automated Western immunoblotting results

AWB yielded 395/602 (66%) negative and 207/602 (34%) positive serum samples (Table 3); giving an 81% sensitivity as 181/223 COVID-19 patients were positive (nucleocapsid detected in 180/223 (76%) and spike in 67/223 (30%), respectively); and a 93% specificity as 26/379 (7%) non-COVID-19 patients were positive; applying above reported cut-off criteria (Figures 1 and 2a). Accordingly, positive (PPV) and negative predictive values (NPV) were of 87% and 89%, respectively. Sera from COVID-19 patients were collected with a median of 13 days (range 0 to 165) after the onset of symptoms. Sensitivity was 54% among sera collected less than 10 days after the onset of symptoms and increased to 94% among sera collected more than 10 days after the onset of symptoms (Figure 3). AWB had a 90% agreement with the herein compared ELISA assay (Cohen’s Kappa=0.64) as the latter was positive in 22/27 (81.5%) COVID-19 patients and 6/170 (3.5%) non-COVID-19 patients, yielding a sensitivity of 81.5% and specificity of 97% (Table 4).

**Table 3.** Results of automated Western immunoblotting including the 602 sera tested.

|  | Negative controls |  |  |  | Sera from patients diagnosed with others HCoV's (n=76) | Sera collected before the pandemic from patients with unknown status for HCoV's (n=266) | Sera from COVID-19 positive patients (n=223) | Total (n=602) |
| --- | --- | --- | --- | --- | --- | --- | --- | --- |
|  | Early sera from COVID-19 positive patients (n=10) | Sera from negative HCoV's patients collected before the pandemic (n=13) | Sera from healthcare workers highly exposed to SARS-CoV-2 (n=14) | All (n=37) |  |  |  |  |
| Positive sera (Nb,%) |  |  |  |  |  |  |  |  |
| Nucleocapside reactivity | 1 (10%) | 0 (0%) | 2 (14) | 3 (8) | 9 (12) | 12 (4.5%) | 180 (81) | 204 (34) |
| Spike reactivity | 0 | 0 | 0 | 0 | 2 (3) | 0 | 67 (30) | 69 (11%) |
| Total | 1 (10%) | 0 (0%) | 2 (14) | 3 (8) | 11 (14.5) | 12 (4.5%) | 181 (81) | 207 (34%) |

**Table 4.**
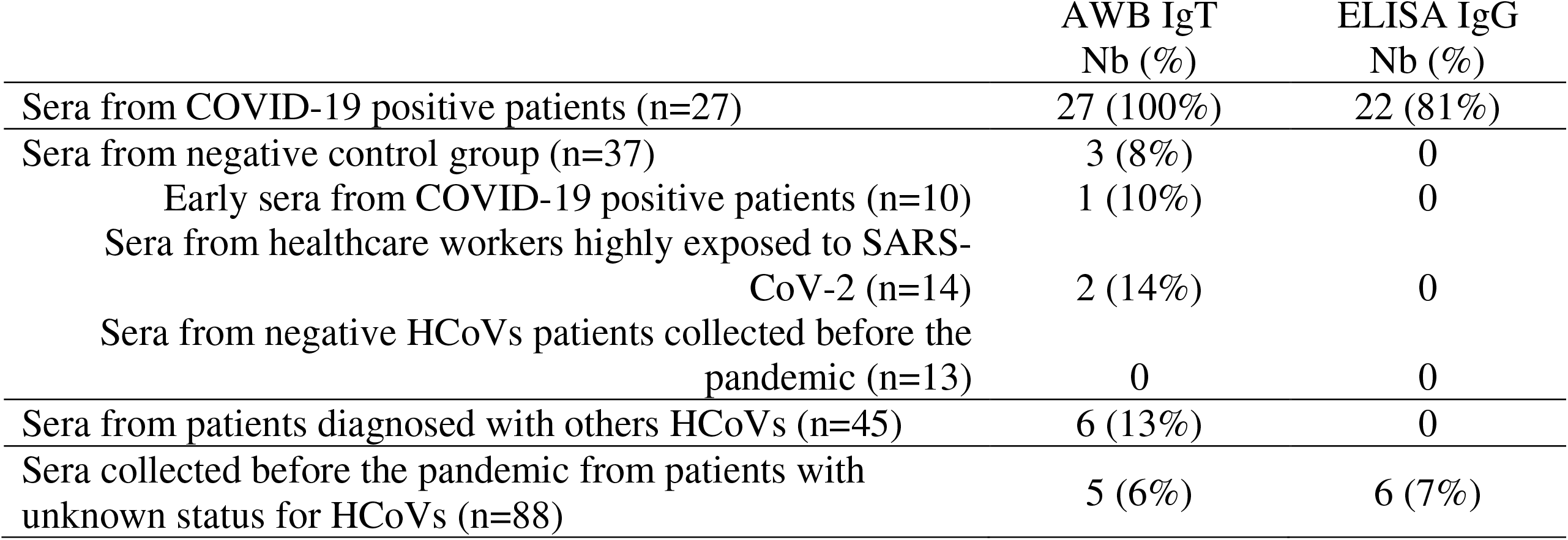
**Comparison between automated Western immunoblotting and commercial SARS-CoV-2 IgG ELISA on 197 sera**.

|  | AWB IgT<br>Nb (%) | ELISA IgG<br>Nb (%) |
| --- | --- | --- |
| Sera from COVID-19 positive patients (n=27) | 27 (100%) | 22 (81%) |
| Sera from negative control group (n=37) | 3 (8%) | 0 |
| Early sera from COVID-19 positive patients (n=10) | 1 (10%) | 0 |
| Sera from healthcare workers highly exposed to SARS-CoV-2 (n=14) | 2 (14%) | 0 |
| Sera from negative HCoV patients collected before the pandemic (n=13) | 0 | 0 |
| Sera from patients diagnosed with others HCoV (n=45) | 6 (13%) | 0 |
| Sera collected before the pandemic from patients with unknown status for HCoV (n=88) | 5 (6%) | 6 (7%) |

**Figure 1.**
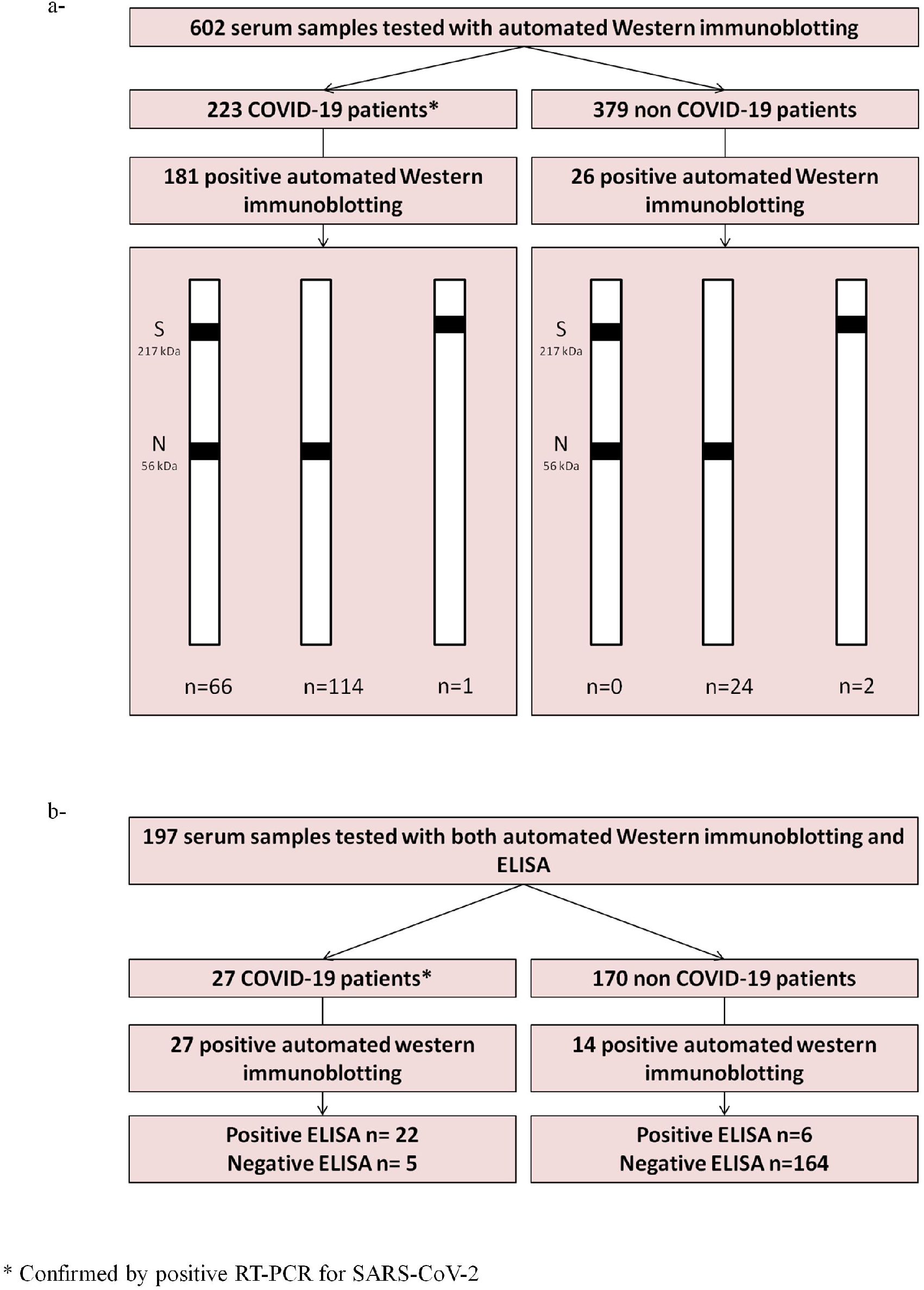
Drawing illustrating (a) overall results of automated Western immunoblotting of 602 sera (b) results comparison with commercially available ELISA in 197 sera.

**Figure 2.**
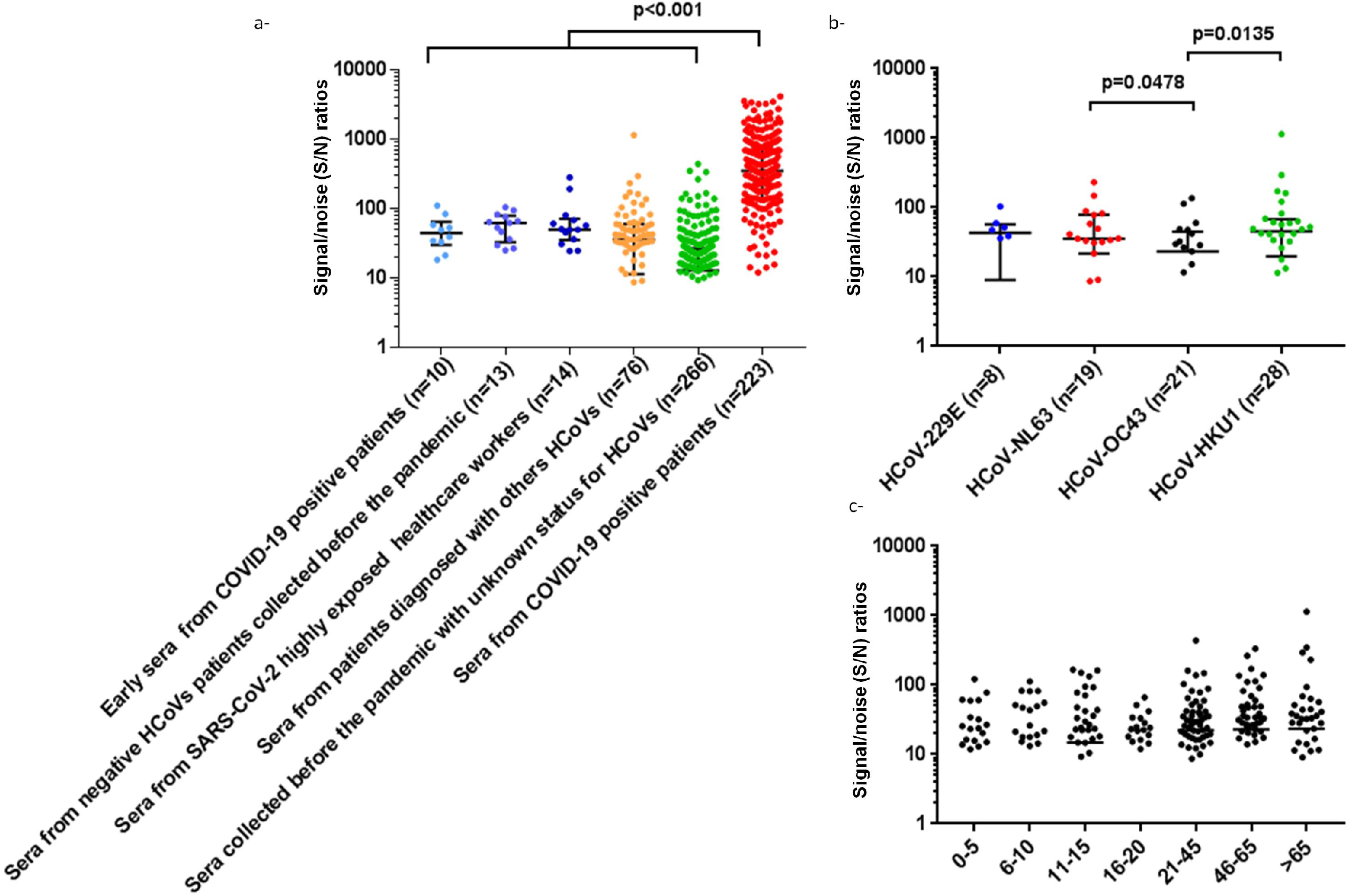
Signal/noise ratio for the detection of nucleocapsid with automated Western immunoblotting: (a) in 602 sera collected from 6 different groups of patients (b) in 76 sera collected from non-COVID-19, HCoVs infected patients (c) in 342 sera collected before the COVID-19 pandemic, classified by age-group.

**Figure 3.**
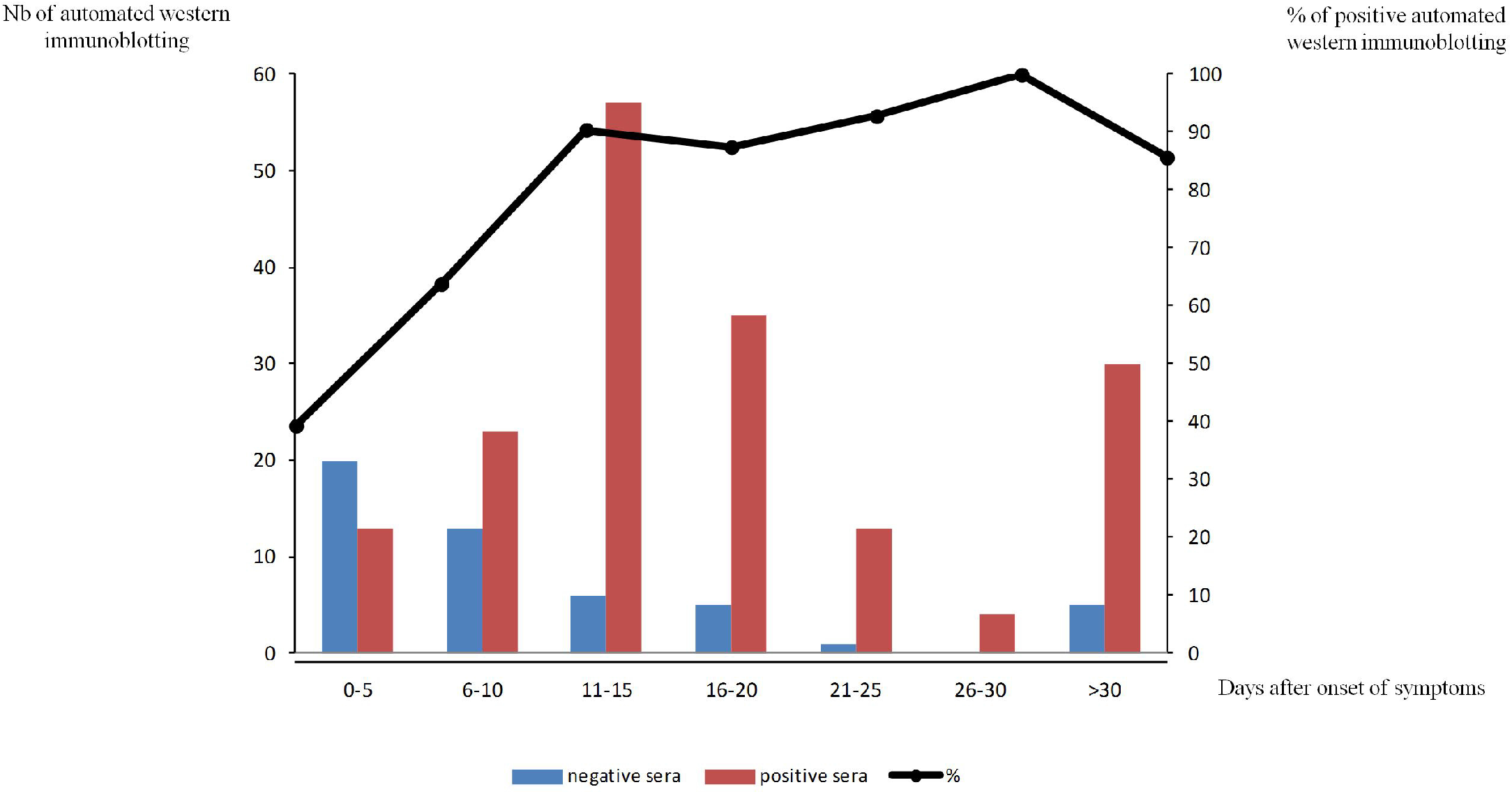
Automated Western immunoblotting detection of anti-SARS-CoV-2 antibodies: positive and negative sera according to delay after the onset of symptoms. The curve represents the proportion of positive sera (%).

Detailing false-positive AWB, antibodies to the nucleocapsid were detected in 3/37 (8%) negative control serum samples. Also, 9/76 (14%) serum sampled from patients diagnosed with seasonal HCoVs, reacted with the nucleocapsid which was detected in 5/28 (18%) of patients with HCoV-HKU1, 2/19 (10.5%) with HCoV-NL63, 2/21 (9.5%) with HCoV-OC43 but in none of HCoV-229E patients (Figure 2b). In addition, one HCoV-HKU1 serum and one HCoV-NL63 serum reacted against the spike protein, increasing the number of total cross-reactions to 11/76 (14.5%) for this group. Among 266 serums sampled before the COVID-19 epidemic in France, albeit of unknown status for HCoVs, 12/266 (4.5%) reacted against the nucleocapsid but none against the spike protein.

Most cross reactivities were detected in 46-65-year-old patients (7/63) and more than 65-year-old (4/43) patients (Figure 2c). Cross reactivity was more prevalent in subjects > 21 years (15/173) than in children ≤ 15 years (6/126) but this difference was not significant (*p* = 0.25, Fisher’s exact test).

## 4. DISCUSSION

An AWB, incorporating whole SARS-CoV-2 viral particles, was demonstrate to be efficient in detecting specific antibodies against SARS-CoV-2 dual nucleocapsid and spike proteins, achieving a 87% PPV and a 89% NPV for COVID-19, in the population tested in this study. Accordingly, dual nucleocapsid and spike protein detections exhibited 81% sensitivity and 93% specificity. Indeed, the spike protein was detected in only two non-COVID-19 patients whereas the nucleocapsid protein was detected in 24 non-COVID-19 patients, including 11 patients diagnosed with HCoVs. In our study, AWB results were consistent with results obtained using a commercially ELISA incorporating recombinant spike-1 protein. The serological observations obtained in this study therefore indicated that it is worth developing next generation serological assays incorporating both the nucleocapsid and the spike proteins, in order to achieve almost 100% sensitivity and 100% specificity of SARS-CoV-2 infection, which is not the situation with first generation, commercially-available serological assays (7, 11, 14).

It should be noted that cross-reactivity was more prevalent in patients infected with other betacoronaviruses (accounting for 31% of cross-reactivity) than in patients infected with alphacoronaviruses (accounting for 12% of cross-reactivity); being mainly supported by SARS-CoV-2 nucleocapsid (in 92% of cross-reacting serum samples); and mostly found in adult patients older than 46 years (accounting for 52% of sera with cross-reactivity). Our observations are consistent with previous reports that cross-reactions were observed with nucleocapsid while serological assays incorporating the spike protein have been reported to be more specific but less sensitive (4–6, 15–18). Cross-reactivity has been described between endemic coronaviruses and SARS-CoV-2 (19). Several studies reported the presence of antibodies reacting with SARS-CoV-2 spike and nucleocapsid proteins in serum sampled before the pandemic and in HCoVs patients (20, 21).

In a few previous reports of the described AWB (22–24), a recombinant protein was used as the antigen whereas we used purified virus antigen directly produced in the biosafety level 3 laboratory (13). This fact could explain in part the important difference of sensitivity for the spike protein compared to the nucleocapsid, in our assay. Thereby, serum dilution was a critical parameter as the spike protein was detected only for a low, 1:2 dilution of serum. Nevertheless, the herein described AWB assay demonstrated a better standardization and reproducibility than conventional Western immunoblotting, proved to be user-friendly and enabled analyzing 24 serum samples in less than 4 hours. Result interpretation was not only based on presence/absence and intensity of bands but a chemiluminescent image was automatically analyzed with software allowing noise reduction. The “virtual image” of reactions present in the capillaries could be represented by peaks on electropherogram or lane views.

In conclusion, the herein described AWB may be incorporated as a first line serological test for the diagnosis of exposure to SARS-CoV-2 if limited series have to be investigated; or as a second-line assay to confirm or exclude the diagnosis of COVID-19 especially in patients with negative, doubtful and discrepant RT-PCR results, and may be used to measure past exposition to the virus.

## Data Availability

All relevant data are made available in the manuscript and supplementary files.

## ACKNOWLEDGMENTS

The authors acknowledge the contribution of the technical staff of the IHU Méditerranée Infection Laboratory. This work was supported by IHU Méditerranée Infection, Marseille, France. RJ benefits from a PhD grant by fondation Mediterranée Infection, Marseille, France.

## FINANCIAL SUPPORT

This study was funded by ANR-15-CE36-0004-01 and by ANR “Investissements d’Avenir” Méditerranée Infection 10-IAHU-03.

## DECLARATION OF COMPETING INTEREST

None to declare.

## Supplementary Figure

**Figure S1**. Strips of conventional western immunoblotting (a) and lane view of automated Western immunoblotting (b) incubated with serum collected from one COVID-19 positive patient quoted “+” and serum collected from one non COVID-19 patient quoted “-”. The first lane represents the molecular mass marker in kDa. (c) chromatogram of chemiluminescence intensity detected by Jess™ Simple Western in the capillaries on positive (blue) and negative (green) sera.

## Notes

### Competing Interest Statement

The authors have declared no competing interest.

### Funding Statement

This study was funded by ANR-15-CE36-0004-01 and by ANR Investissements d Avenir Mediterranee Infection 10-IAHU-03

### Author Declarations

All sera were retrospectively tested and no sample was collected specifically for this study which was approved by our institution s ethics committee under No.2020-024.

